# Digitally managing depression: a fully remote randomized attention-placebo controlled trial

**DOI:** 10.1101/2023.04.05.23288184

**Authors:** Aaron Kandola, Kyra Edwards, Marie AE Muller, Bettina Dührkoop, Bettina Hein, Joris Straatman, Joseph F Hayes

## Abstract

**Background:** Depression is a common and disabling condition. Digital apps may augment or facilitate care, particularly in under-served populations. We tested the efficacy of juli, a digital self-management app for depression in a fully remote randomized controlled trial.

**Methods:** We completed a pragmatic single-blind trial of juli for depression. We included participants aged over 18 who self-identified as having depression and scored 5 or more on the Patient Health Questionnaire-8 (PHQ-8). Participants were randomly assigned (1:1) to receive juli for 8 weeks or a limited attention-placebo control version of the app. Our primary outcome was the difference in PHQ-8 scores at 8 weeks. Secondary outcomes were remission, minimal clinically important difference, worsening of depression, and health-related quality of life. Analyses were per protocol (primary) and modified intention-to-treat (secondary). The trial was registered at the ISRCTN registry (ISRCTN12329547).

**Results:** Between May 2021 and January 2023, we randomised 908 participants. 662 completed the week 2 outcome assessment and were included in the modified intention-to-treat analysis, and 456 completed the week 8 outcome assessments (per protocol). The mean baseline PHQ-8 score was consistent with a diagnosis of moderately severe depression. In the per-protocol analysis, the juli group had a lower mean PHQ-8 score (10.78, standard deviation 6.26) than the control group (11.88, standard deviation 5.73) by week 8 (baseline adjusted β-coefficient -0.94, 95%CI -1.87 to -0.22, p=0.045). Remission and minimal clinically important difference were increased in the juli group at 8 weeks (adjusted odds ratio 2.22, 95%CI 1.45-3.39, p<0.001 and adjusted odds ratio 1.56, 95%CI 1.08 to 2.27, p=0.018). There were no between-group differences in health-related quality of life physical or mental component scores or worsening of depression.

**Conclusion:** Use of juli reduced symptoms of depression at 8 weeks compared with an attention-placebo control. The juli app is a digital self-management tool that could increase accessibility of evidence-based depression treatments.

## Introduction

Depression is a major contributor to the global burden of disease. Each year, millions of people are diagnosed with depression, and is likely to be the leading cause of disability in high-income countries by 2030^1^. The mainstay of treatment for depression includes psychological intervention and antidepressant medication. Barriers to accessing care include high costs, low availability, long waiting lists and stigma. Digital interventions may remove some of these barriers by offering scalable solutions that are convenient and timely for people with depression^2^.

Recent meta-analyses of randomised controlled trials (RCTs) of smartphone app-based psychological interventions for depression symptoms find a small to moderate reduction in symptoms compared to placebo, such as waiting list control^3,4^. However, subgroup analyses find no difference in depressive symptoms in studies with active control groups, which is corroborated by the findings of other systematic reviews^5^. Reviews also only include small numbers of trials, which are frequently of short duration and/or include few participants. Participants often do not have severe depression^3^ Many of the apps included in the reviews are no longer (or have never been) commercially available, highlighting that translation from research to clinical impact is difficult in this space. Other challenges include low levels of retention and engagement with digital health apps in trials and real-world use^6^. The reviews all conclude that further methodologically robust RCTs are required. We sought to address some of the limitations of previous RCTs by pre-registering our trial, powering it adequately to detect between-group differences, following-up users for an adequate duration, efforts to minimise attrition and avoiding recruitment from unrepresentative populations (such as from mood disorders clinics). Our RCT was fully remote, increasing cost-effectiveness, time efficiency and reach.

The digital health app juli, aims to support people with depression via numerous evidence-based approaches, including mood tracking, medication reminders, positive affect journaling, data visualisation of sleep, activity, exercise, heart rate variability and behavioural activation recommendations about how to improve these parameters^7^. As such, it combines many of the elements that have been found to be effective in research-grade apps for depression. However, there has been less evaluation of consumer-grade apps in real-life practice. This is important as many popular health apps have not been scrutinised in the way new interventions traditionally would be. We hypothesised that individuals randomized to juli would have a greater reduction in depression symptoms at 8 weeks than those in an attention placebo control group.

## Methods

### Study design and participants

We conducted a fully remote pragmatic single-blind, placebo control randomised controlled trial of people with depression from anywhere in the world. Individuals we eligible for inclusion if they were aged 18 to 65, were English speaking, had access to an iPhone, and self-identified as having depression, with a score of 5 or more on the Patient Health Questionnaire 8-item version (PHQ-8) at baseline. We recruited participants via online adverts, social media posts and self-help groups for depression. Recruitment was from May 2021 until January 2023. All participants provided written informed consent via a consent form within the app. Ethical approval was from the University College London Ethics Committee (ID number 19413/001). The trial was registered on the ISRCTN registry (https://doi.org/10.1186/ISRCTN12329547).

### Randomisation and masking

We randomly assigned participants (1:1) to a full version of juli or an attention-placebo control version. Block randomisation was conducted within the app using automated code, with random block sizes between 4 and 8. The researchers and independent statisticians were masked to treatment allocation until the completion of the analysis.

### Procedures

Participants were either allocated to the full version of the juli app or an attention placebo control app group. If allocated to the full version of the app, participants were prompted to open the app each day via an automated alert at the time of their choosing. They were asked to rate how they were feeling on a scale using 5 emoji faces and a circumplex model with mood on the x-axis and energy on the y-axis^8^. Individuals were also able to track things that they considered as important contributors to their mood^9^. The app passively gathered information via smartphone and smartwatch sensors on sleep, activity, workouts, menstrual cycle, and heart rate variability on a daily basis and presented this data to the participant, showing associations with mood^10^. The app then provided recommendations about these parameters to guide healthy behaviours via behavioural activation^11^. The app includes a medication reminder function that can be set by the participants to improve medication adherence^12^. Participants were also encouraged to engage in positive affect journaling via the app (Figure 1)^13^. The juli app was designed by a psychiatrist and experts in gamification^14^. It is grounded in evidence-based treatments. Participants were guided towards all elements of the app, but did not have to engage with elements they did not like. If allocated to the control arm, participants were prompted to open the app each day via an automated alert and to rate how they were feeling on a scale using 5 emoji faces. Control participants did not receive any further intervention.

**Figure 1.**
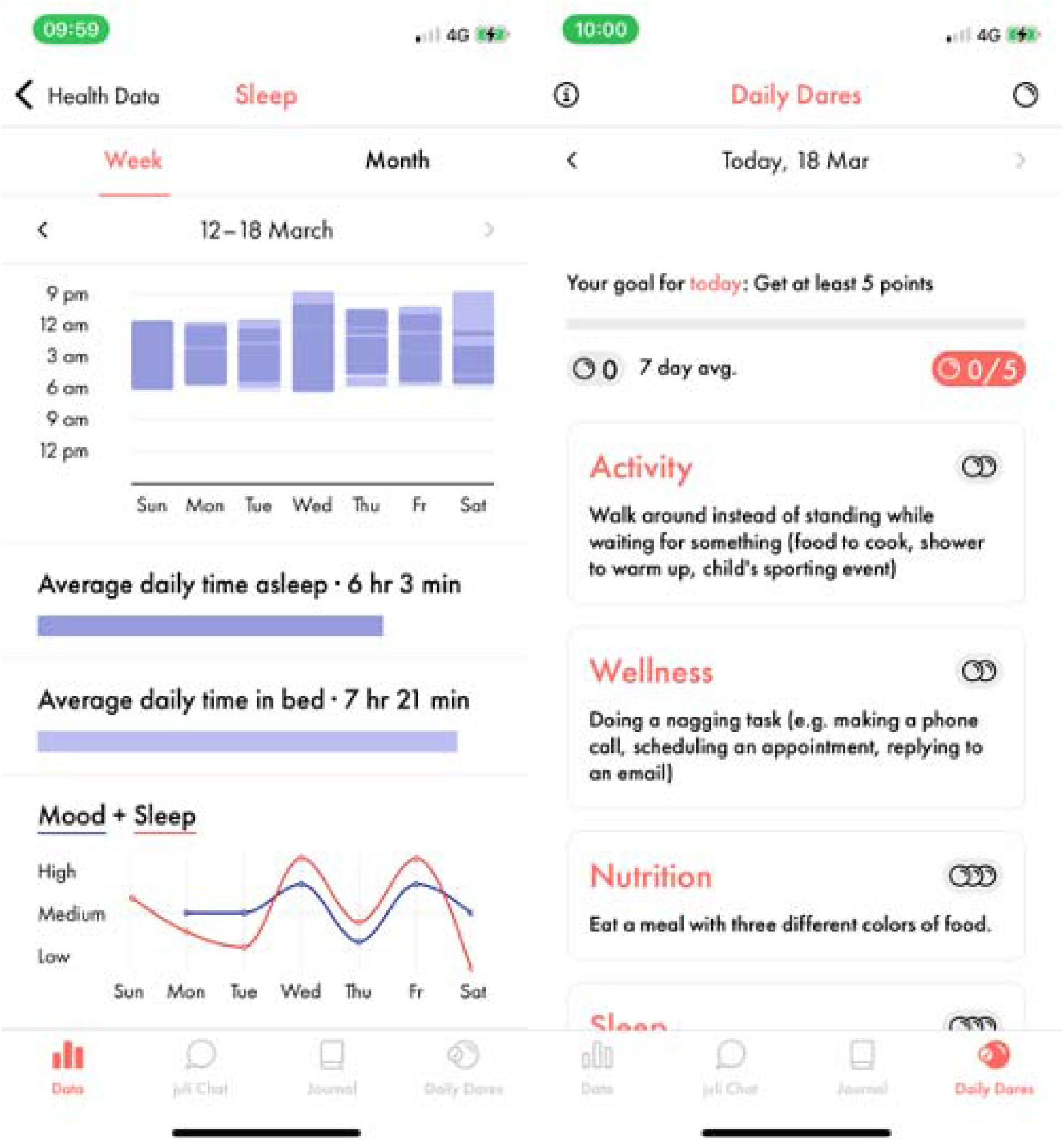
Example juli screen shots.

Baseline assessments and follow-up assessments at two, four, six and eight weeks were all completed within the app. Participants completed the PHQ-8 for depression symptoms and the 12-Item Short Form Health Survey (SF-12) for health-related quality of life at baseline. The PHQ is a widely used self-completed depression scale that is recommended by the Common Measures in Mental Health Science Governance board as one of a core list of research questionnaires that should be used by funded researchers. The PHQ-9 closely matches the DSM-IV criteria for a major depressive episode and may be more sensitive to change than other measures of depression, such as the Hamilton Rating Scale for Depression and the Beck Depression Inventory^15^. The PHQ-8 excludes the question about suicidality, which is preferred in studies where patient contact is remote, such as via digital technologies or telephone. Research indicates that the deletion of this question has little effect on the scale’s psychometric properties because this question is the least frequently endorsed item on the PHQ-9. Subsequently, the PHQ-8 has identical scoring thresholds for depression severity, with higher scores representing more severe depression^16,17^. The SF-12 is a self-reported health status measure^18^. Possible scores range from 0 to 100, with higher scores indicating better quality of life.

### Outcomes

The primary outcome was the total score on the PHQ-8 at 8 weeks.

The secondary outcomes were: 1) PHQ-8 score as a continuous outcome at 2, 4, 6 and 8 weeks in a repeated measures analysis, 2) PHQ-8 score as a binary outcome where remission is a score of <10 at 8 weeks, 3) remission at 2, 4, 6 and 8 weeks in a repeated measures analysis, 4) Difference in SF-12 physical and mental component scores at 8 weeks and, 5) SF-12 physical and mental component scores at 4 and 8 weeks in a repeated measures analysis.

We added post-hoc outcomes that included: 1) achieving a minimal clinically important difference (MCID) at 8 weeks defined by the effective dose 50 method, which accounts for baseline severity and is the smallest difference in PHQ-8 scores that are of perceived benefit^19^, 2) a worsening of depression, defined as a >20% increase in PHQ-8 from baseline.

### Statistical analyses

Our analysis plan was pre-printed (https://discovery.ucl.ac.uk/id/eprint/10129350/) and included on the ISRCTN registry. We followed the Consolidated Standards of Reporting Trials (CONSORT) guidelines in reporting and analysing our data ^17^. Primary and secondary outcomes were described in the published protocol and on the ISRCTN registry before the study started.

The best estimate of a MCID in PHQ-8 is between 11% and 14%, with a standard deviation of 0.32-0.38^20,21^. 80% power at the two-sided 5% significance level requires a total sample size of 378. Allowing for 26% attrition^22^, we aimed to recruit 238 participants per arm. Power calculations were carried out using Stata.

The primary outcome was the difference in total PHQ-8 score at 8 weeks between control and intervention groups in a per protocol analysis. This was estimated with a linear regression model adjusted for baseline PHQ-8. We calculated the odds ratio of remission at 8 weeks (PHQ-8<10), achieving MCID and worsening of depression, adjusting for baseline severity using logistic regression. Repeat measures analyses were completed using linear or logistic mixed effect models adjusting for baseline severity.

We also examined all outcomes in a modified intention-to-treat analysis including all randomised participants with a complete baseline and week 2 PHQ-8. Missing outcome data were imputed by last observation carried forward and controlled multiple imputation^23^.

All analyses were completed by independent statisticians using Stata and R, who have no financial conflict of interest with the company providing the juli app.

## Results

### Baseline characteristics

We recruited 456 individuals who were retained in the trial for 8 weeks and formed the basis of our primary per protocol analysis (Figure 2). The majority of participants were female and had experienced depression for more than five years (Table 1). The majority were diagnosed by a physician and continued to be in regular or occasional contact with a doctor about their depression. The mean PHQ-8 score at baseline was 16.16 (standard deviation (SD) 4.71).

**Table 1.** Baseline characteristics.

|  | Intervention<br>(n=200) | Control<br>(n=256) | All<br>(n=456) |
| --- | --- | --- | --- |
| Age | 34.39 (12.41) | 32.65 (12.28) | 33.41 (12.35) |
| Gender |  |  |  |
| Female | 154 (77.00) | 202 (78.91) | 356 (78.07) |
| Male | 33 (16.50) | 37 (14.45) | 72 (15.69) |
| Other | 13 (6.50) | 17 (6.64) | 30 (6.58) |
| Depression duration |  |  |  |
| <1 month | 1 (0.50) | 1 (0.39) | 2 (0.44) |
| 1 to <3 months | 7 (3.50) | 2 (0.78) | 9 (1.97) |
| 3 months to <1 year | 2 (1.00) | 14 (5.47) | 16 (3.51) |
| 1 year to <2 years | 11 (5.50) | 15 (5.86) | 26 (5.70) |
| 2 years to <5 years | 43 (21.50) | 49 (19.14) | 92 (20.18) |
| >5 years | 136 (68.20) | 175 (68.36) | 311 (68.20) |
| Physician contact |  |  |  |
| Regular | 76 (38.00) | 112 (43.75) | 188 (41.23) |
| Occasional | 67 (33.50) | 64 (25.00) | 131 (28.73) |
| Not anymore | 32 (16.00) | 38 (14.84) | 70 (15.35) |
| Never | 25 (12.50) | 42 (16.41) | 67 (14.69) |
| Diagnosed by a physician |  |  |  |
| Yes | 179 (88.61) | 219 (85.55) | 397 (87.06) |
| No | 22 (11.00) | 37 (14.45) | 59 (12.94) |
| PHQ-8 total score<sup>1</sup> | 16.09 (4.94) | 16.30 (4.67) | 16.21 (4.78) |
| SF-12 physical health subscale<sup>2</sup> | 46.61 (9.29) | 45.05 (9.81) | 45.74 (9.61) |
| SF-12 mental health subscale<sup>3</sup> | 22.15 (7.87) | 22.09 (7.70) | 22.12 (7.79) |

Data are n (%) or mean (SD). <sup>1</sup>Patient Health Questionnaire, 8-item version (possible range 0–24), <sup>2</sup>Short-Form Health Survey12 physical health subscale (possible range 0–100), <sup>3</sup>Short-Form Health Survey-12 mental health subscale (possible range 0–100). Data used in per protocol analysis of individuals completing week 8 PHQ-8

**Figure 2.**
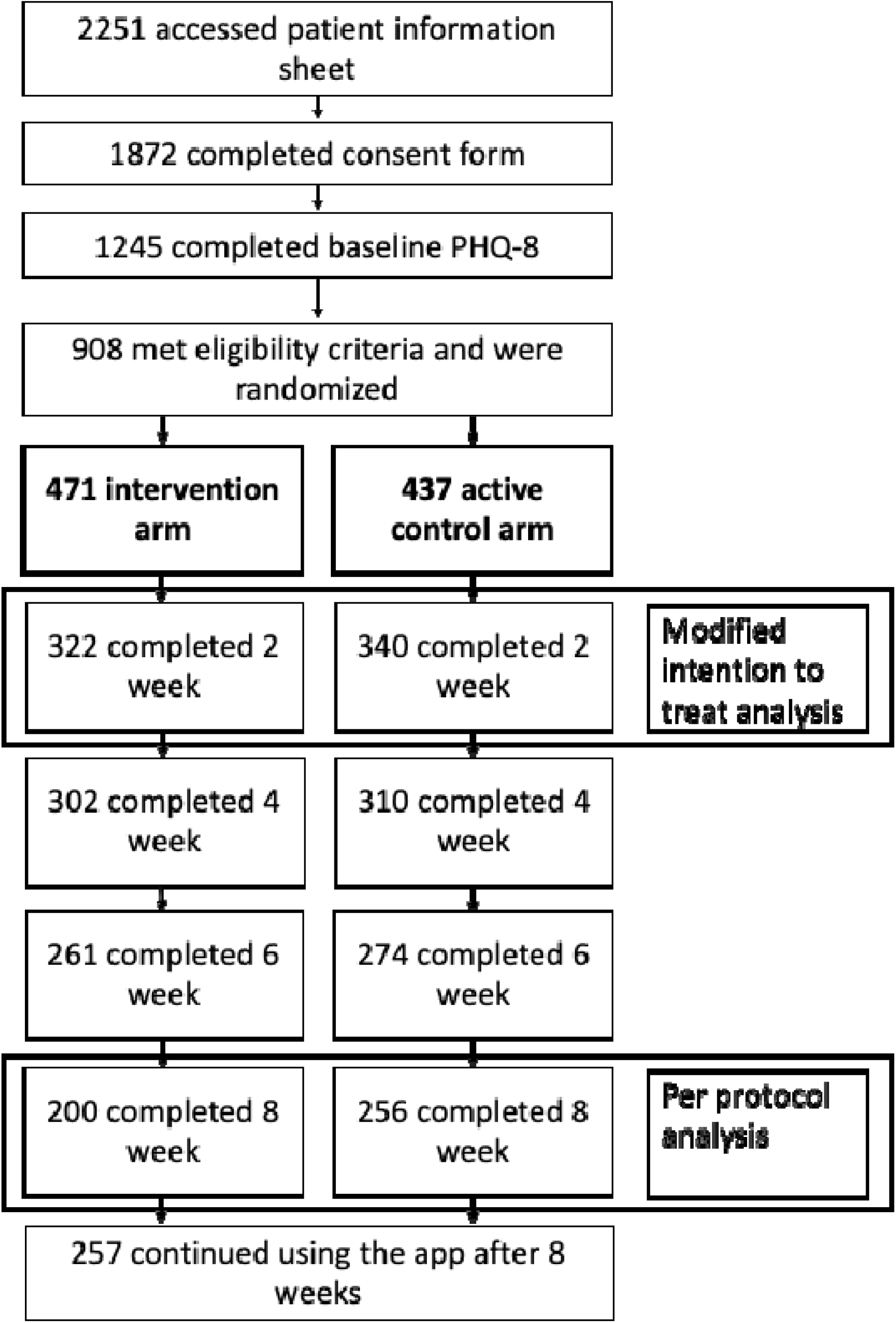
CONSORT diagram.

### Per protocol analysis

At 8 weeks, participants in the intervention group had a mean PHQ-8 score of 10.78 (SD 6.26) and participants in the control group had a mean of 11.88 (SD 5.73). After accounting for baseline PHQ-8 score the intervention group had lower depression symptom scores at 8 weeks (−0.94, 95% confidence interval (CI) -1.87 to -0.22, p=0.045). The odds of being in remission by week 8 was higher in the intervention group after accounting for baseline depression severity (adjusted odds ratio 2.22, 95%CI 1.45 to 3.39, p<0.001). Participants in the intervention group were more likely to experience MCID (adjusted odds ratio 1.56, 95%CI 1.08 to 2.27, p=0.018) than those in the control group. Repeat measures analyses of these outcomes at 2, 4, 6 and 8 weeks suggest that this effect was maintained over time (Supplement Table 2). We found no effect of the intervention on SF-12 mental or physical component scores. The odds of experiencing worsening symptoms were similar in the intervention and control groups (adjusted odds ratio 0.83, 95%CI 0.38 to 1.81, p=0.633).

### Intention-to-treat analysis

Modified intention-to-treat analysis included 322 participants in the intervention group and 340 controls who completed baseline and week 2 PHQ-8 (Supplement Table 1). Baseline characteristics of participants in these groups were similar to the per protocol analysis. In the last observation carried forward dataset, there was no clear difference between intervention and control groups after accounting for baseline severity (β-coefficient -0.72, 95%CI -1.49 to 0.06)(Supplement table 2) at week 8. However, the repeated measures analysis of PHQ-8 scores found a positive effect of the intervention on depression symptoms. The odds ratios for remission at 8 weeks and remission in repeated measures were similar to the per protocol analysis, and suggested higher odds of remission in the intervention group. The odds ratios for MCID and worsening of symptoms were consistent with the per protocol results. Results from the multiple imputation dataset were consistent with the last observation carried forward findings.

## Discussion

We found a small reduction in depression symptoms in participants using juli for 8 weeks compared to an attention-placebo control. Both groups had a reduction in PHQ-8 scores over the course the trial. The mean intervention arm PHQ-8 score decreased by 37%, a reduction considerably greater than the MCID^24^. Participants allocated to juli were more than twice as likely to be in remission by 8 weeks, and more likely meet the threshold for MCID in depression symptoms.

The participants had a mean baseline PHQ-8 score consistent with a diagnosis of moderately severe depression, the majority had longstanding depression and were under the ongoing care of a physician. Therefore these participants were chronically unwell and potentially experiencing depressive symptoms that were difficult to treat. This is important as our participants differ from those included in many digital depression intervention RCTs in terms of severity and duration^3^.

The improvement in the attention-control group was of a similar magnitude to other placebo controlled trials of depression interventions^25^. In addition, even basic mood monitoring, as required by our control group, has been found to decrease depression symptoms^26^. This may have reduced difference observed between intervention and control groups, compared to an inactive control (such as waiting list).

Findings from meta-analyses support the use of digital technologies for the treatment of depression. However, many of the interventions reviewed are not available to patients, often because they are not available commercially or via healthcare providers. juli uses a combination of evidence-based approaches to support symptom reduction in depression and is available globally on Apple and Android formats. Our trial provides methodologically robust evidence of efficacy.

### Strengths and limitations

Our RCT has a number of strengths and limitations. We successfully recruited, screened, randomized, treated, and assessed a geographically dispersed sample of participants. We modified the juli app so that for RCT participants could consent, be randomized and take the baseline assessment within the app. This facilitated global recruitment, with low cost, in a pragmatic manner, with good external validity. However, this also meant that we lacked information on potentially important baseline characteristics, such as social determinants of health, as we did not want to overburden the participants. Despite this, balance in the recorded baseline characteristics after randomisation supports the assumption that randomisation was successful. Additionally, we did not include a large battery of outcome measures, which may have shed further light on our findings. For example, improvements in anxiety symptoms have been found in RCTs of interventions for depression^21^.

We pre-registered our analysis plan and did not deviate from this. Attrition was higher than we anticipated (49.78% from randomisation to week 8). The majority of the attrition occurred between randomisation and week 2, which is common in RCTs, including for depression apps. A recent meta-analysis of dropout rates found similar attrition after adjusting for publication bias^22^. We overcame this by recruiting until we had sufficient numbers who had completed the week 8 outcome measures and examined differences in completers vs non-completers.

We analysed participants using the app for 8 weeks in our primary analysis to focus on the effects of maintained use. Baseline characteristics were similar in the modified intention-to-treat and primary analysis, and results were consistent, suggesting little difference between completers and non-completers. Following the publication of our protocol and commencement of our RCT, newer research on the PHQ-8 MCID was published suggesting that we may have underestimated our power calculations^19,24^. We used these newer methods to derive a post hoc MCID outcome^19^. We used two imputation methods for the intention-to-treat analysis that make different assumptions^23^. Results from both methods did not differ.

We found that most participants were diagnosed by a doctor and remained under their care. This highlights the severity of our participants depression, but does not suggest that people were accessing juli because of an absence of traditional care. Individuals with no previous or ongoing healthcare may differentially benefit from digital technologies. Digital apps could be one solution to an overburdened healthcare system, particularly in groups or areas where accessing treatment for mental health conditions is challenging or stigmatising^27^. However, some people may still be unwilling or unable to use apps for health, despite their relative ease of access. The majority of participants were female, which reflects established differences in sex-specific rates of depression and help-seeking for depression^28^. Some of our participants (∼7%) identified as transgender or non-binary. This group is under-represented in research but has higher risk of depression and other mental disorders^29^. Our high uptake suggests that digital technologies may be a better way of engaging these populations.

An even bigger problem than the high attrition in RCTs of digital apps for depression is their lack of real-world retention and engagement. The proportion of 30-day retention is less than 10% across mental health apps^6^. The 30-day retention of juli users (non-RCT participants) is 25%. This highlights that engagement and the potential clinical benefit can be increased by methods employed by juli. However, it is unclear which specific features increase engagement. For example, a 2021 meta-analysis found no benefit of gamification^30^. More research is needed to understand which depression app features are integral to improving mental health symptoms.

## Conclusion

The juli app can reduce depressive symptoms within 8 weeks, with increased probability of remission and MCID. As such it represents a low-risk addition to the care package of people with mild to severe depression. Further research is required to determine the most cost-effective technical support processes to enhance engagement, and how juli could be implemented in current public health or clinical care models.

## Data Availability

All data produced in the present study are available upon reasonable request to the authors.

## Author Contributions

JFH conceived the study; JFH, AK, BD, BH and JS designed the study; AK, BD and BH collected the data; KE and MM analysed the data; JFH wrote the initial draft; all authors edited and approved the final manuscript.

## Acknowledgements

AK is supported by the UK Research and Innovation (UKRI) Digital Youth Programme award [MRC project reference MR/W002450/1], which is part of the AHRC/ESRC/MRC Adolescence, Mental Health and the Developing Mind programme. JFH is supported by the UK Research and Innovation grant MR/V023373/1, the University College London Hospitals NIHR Biomedical Research Centre, and the NIHR North Thames Applied Research Collaboration.

## Data Availability

All data produced in the present study are available upon reasonable request to the authors.

