## Supplemental material for "Digitally managing depression: a fully remote randomized attention-placebo controlled trial"

**Supplement table 1. Baseline characteristics of individuals in modified intention-to-treat analysis**

|  | Intervention  (n=322) | Control  (n=340) | All  (n=662) |
| --- | --- | --- | --- |
| Age | 33.92 (12.10) | 31.75 (12.24) | 32.80 (12.21) |
| Gender |  |  |  |
| Female | 249 (77.33) | 272 (80.00) | 521 (78.70) |
| Male | 52 (16.15) | 48 (14.12) | 100 (15.11) |
| Other | 21 (6.52) | 20 (5.88) | 41 (6.19) |
| Depression duration |  |  |  |
| <1 month | 1 (0.31) | 1 (0.29) | 2 (0.30) |
| 1 to <3 months | 13 (4.04) | 3 (0.88) | 16 (2.42 |
| 3 months to <1 year | 5 (1.55) | 23 (6.76) | 28 (4.23) |
| 1 year to <2 years | 29 (9.01) | 29 (8.53) | 58 (8.76) |
| 2 years to <5 years | 68 (21.12) | 65 (19.12) | 133 (20.09) |
| >5 years | 206 (63.98) | 219 (64.41) | 425 (64.20) |
| Physician contact |  |  |  |
| Regular | 114 (35.40) | 148 (43.53) | 262 (39.58) |
| Occasional | 109 (33.85) | 84 (24.71) | 193 (29.15) |
| Not anymore | 51 (15.84) | 49 (14.41) | 100 (15.11) |
| Never | 48 (14.91) | 59 (17.35) | 107 (16.16 |
| Diagnosed by a physician |  |  |  |
| Yes | 281 (87.27) | 289 (85.00) | 570 (86.10) |
| No | 41 (12.71) | 51 (15.00) | 92 (13.90) |
| PHQ-8 total score^1^ | 16.08 (4.87) | 16.24 (4.56) | 16.16 (4.71) |
| SF-12 physical health subscale^2^ | 46.78 (9.33) | 45.28 (9.77) | 46.01 (9.58) |
| SF-12 mental health subscale^3^ | 22.05 (7.88) | 21.98 (8.06) | 22.02 (7.97) |

Data are n (%) or mean (SD). ^1^Patient Health Questionnaire, 8-item version (possible range 0–24), ^2^Short-Form Health Survey-12 physical health subscale (possible range 0–100), ^3^Short-Form Health Survey-12 mental health subscale (possible range 0–100).

**Supplement table 2. Outcomes**

|  | **Effect estimate** | **p-value** |
| --- | --- | --- |
| **Per protocol** |  |  |
| PHQ-8 at 8 weeks^1^ | -0.94 (-1.87 to -0.02) | 0.045 |
| Remission at 8 weeks^2^ | 2.22 (1.45 to 3.39) | <0.001 |
| PHQ-8 repeated measures (2, 4, 6, 8 weeks)^1^ | -0.81 (-0.51 to -0.12) | 0.022 |
| Remission repeated measures (2, 4, 6, 8 weeks)^2^ | 2.27 (1.41-3.64) | 0.001 |
| SF-12 physical component score at 8 weeks^1^ | -0.07 (-1.26 to 1.12) | 0.911 |
| SF-12 mental component score at 8 weeks^1^ | 1.38 (-0.13 to 2.90) | 0.074 |
| SF-12 physical component score repeated measures (4, 8 weeks)^1^ | 0.30 (-0.70 to 1.30) | 0.561 |
| SF-12 mental component score repeated measures (4, 8 weeks)^1^ | 0.80 (-0.42 to 2.01) | 0.199 |
| MCID at 8 weeks^2^ | 1.56 (1.08 to 2.27) | 0.018 |
| Worse at 8 weeks^2^ | 0.83 (0.38 to 1.81) | 0.633 |
| **Intention to treat, last observation carried forward** |  |  |
| PHQ-8 at 8 weeks^1^ | -0.72 (-1.49 to 0.06) | 0.073 |
| PHQ-8 at 2 weeks^1^ | -1.01 (-1.66 to -0.37) | 0.021 |
| PHQ-8 at 4 weeks^1^ | -0.85 (-1.60 to -0.10) | 0.027 |
| PHQ-8 at 6 weeks^1^ | -0.74 (-1.48 to -0.01) | 0.048 |
| Remission at 8 weeks^2^ | 1.77 (1.26 to 2.51) | 0.001 |
| PHQ-8 repeated measures (2, 4, 6, 8 weeks)^1^ | -0.83 (-1.44 to -0.21) | 0.008 |
| Remission repeated measures (2, 4, 6, 8 weeks)^2^ | 2.23 (1.41 to 3.51) | 0.001 |
| SF-12 physical component score at 8 weeks^1^ | 0.01 (-0.98 to 0.99) | 0.992 |
| SF-12 mental component score at 8 weeks^1^ | 0.58 (-0.68 to 1.84) | 0.367 |
| SF-12 physical component score repeated measures (4, 8 weeks)^1^ | 0.35 (-0.54 to 1.24) | 0.436 |
| SF-12 mental component score repeated measures (4, 8 weeks)^1^ | 0.26 (-0.82 to 1.35) | 0.635 |
| MCID at 8 weeks^2^ | 1.47 (1.08-1.99) | 0.013 |
| Worse at 8 weeks^2^ | 0.85 (0.48 to 1.51) | 0.576 |
| **Intention to treat, controlled multiple imputation** |  |  |
| PHQ-8 at 8 weeks^1^ | -0.78 (-1.61 to 0.04) | 0.063 |
| PHQ-8 at 2 weeks1 | -1.01 (-1.66 to -0.37) | 0.021 |
| PHQ-8 at 4 weeks1 | -0.87 (-1.62 to -0.12) | 0.023 |
| PHQ-8 at 6 weeks1 | -0.65 (-1.42 to 0.12) | 0.097 |
| Remission at 8 weeks^2^ | 1.94 (1.32 to 2.83) | 0.001 |
| PHQ-8 repeated measures (2, 4, 6, 8 weeks)^1^ | -0.65 (-1.21 to 0.09) | 0.022 |
| Remission repeated measures (2, 4, 6, 8 weeks)^2^ | 1.73 (1.21 to 2.47) | 0.003 |
| SF-12 physical component score at 8 weeks^1^ | 0.81 (-0.67 to 2.29) | 0.284 |
| SF-12 mental component score at 8 weeks^1^ | 1.21 (-0.29 to 2.72) | 0.115 |
| SF-12 physical component score repeated measures (4, 8 weeks)^1^ | 1.18 (-0.11 to 2.47) | 0.074 |
| SF-12 mental component score repeated measures (4, 8 weeks)^1^ | 0.44 (-0.74 to 1.61) | 0.466 |
| MCID at 8 weeks^2^ | 1.57 (1.12 to 2.20) | 0.009 |
| Worse at 8 weeks^2^ | 0.89 (0.46 to 1.72) | 0.722 |

^1^β-coefficient, ^2^odds ratio
